# Elevated T cell repertoire diversity is associated with progression of lung squamous cell premalignant lesions

**DOI:** 10.1101/2021.02.25.21252467

**Authors:** Asaf Maoz, Carter Merenstein, Yusuke Koga, Austin Potter, Adam C. Gower, Gang Liu, Sherry Zhang, Hanqiao Liu, Christopher Stevenson, Avrum Spira, Mary E. Reid, Joshua D. Campbell, Sarah A. Mazzilli, Marc E. Lenburg, Jennifer E. Beane

**Author notes:** These authors contributed equally to this work (Asaf Maoz, Carter Merenstein).

## Abstract

**Objective:** The immune response to invasive carcinoma has been the focus of published work, but little is known about the adaptive immune response to bronchial premalignant lesions (PMLs), precursors of lung squamous cell carcinoma. This study was designed to characterize the T cell receptor (TCR) repertoire in PMLs and its association with clinical, pathological, and molecular features.

**Methods:** Endobronchial biopsies (n=295) and brushings (n=137) from high-risk subjects (n=50), undergoing lung cancer screening at approximately 1-year intervals via autofluorescence bronchoscopy and CT, were profiled by RNA-seq. We applied the TCR Repertoire Utilities for Solid Tissue/Tumor (TRUST) tool to the RNA-seq data to identify TCR CDR3 sequences across all samples. In the biopsies, we measured the correlation of TCR diversity with previously derived immune-associated PML transcriptional signatures and outcome. We also quantified the spatial and temporal distribution of shared and clonally expanded TCRs. Using the biopsies and brushes, the ratio of private (i.e., found in one patient only) and public (i.e., found in two or more patients) TCRs was quantified and the CDR3 sequences were compared to those found in curated databases with known antigen specificities.

**Results:** We detected 39,303 unique TCR sequences across all samples. In PML biopsies, TCR diversity was negatively associated with a transcriptional signature of T-cell mediated immune activation (Spearman’s rho −0.34, p < 0.001) associated with PML outcome.

Additionally, in lesions of the proliferative molecular subtype, TCR diversity was decreased in regressive versus progressive/persistent PMLs (p=0.045). Within each patient, TCRs were more likely to be shared between biopsies sampled at the same timepoint than biopsies sampled at the same anatomic location at different times. Clonally expanded TCRs, within a biopsied lesion, were more likely to be expanded at future time points than non-expanded clones. The majority of TCR sequences were found in a single sample, with only 3,396 (8.6%) found in more than one sample and 1,057 (2.7%) found in two or more patients (i.e. public), however, when compared to a public database of CDR3 sequences, 4,543 (11.6%) of TCRs were identified as public. TCRs with known antigen specificities were enriched among Public TCRs (p < 0.001).

**Conclusions:** Decreased TCR diversity may reflect nascent immune responses that contribute to PML elimination. Further studies are needed to explore the potential for immunoprevention of PMLs.

## Introduction

Lung cancer is the second most common cancer in the United States and accounts for approximately one in four cancer deaths[1]. Patients with lung cancer have poor overall survival, in part because the disease is diagnosed after it has already spread to distant sites[1]. Squamous cell lung cancer, the second most common form of lung cancer, is thought to arise through accumulation of genomic and epigenomic alterations to the airway epithelium. These alterations lead to a series of histological changes, ranging from hyperplasia, metaplasia, several grades of dysplasia, carcinoma in situ, and to invasive carcinoma[2,3]. Persistent bronchial dysplasia has been associated with increased risk of incident invasive carcinoma, both at the site of the dysplasia and at other locations in the lung[3,4]. Since squamous cell lung cancer arises through a series of bronchial premalignant lesions (PMLs) that can be evaluated by bronchoscopy, it has been proposed as a model for cancer chemo- and immunoprevention[5–8].

By transcriptionally profiling endobronchial biopsies of PMLs obtained from high-risk smokers undergoing lung cancer screening, we have identified distinct molecular subtypes of squamous PMLs[9]. A proliferative molecular subtype of squamous PML is enriched with bronchial dysplasia. Proliferative subtype PMLs that progressed or persisted were depleted of innate and adaptive immune cells and demonstrated decreased expression of interferon signaling and antigen presentation pathways. Additional studies have shown that high-grade or progressive/persistent PMLs are associated with an immunosuppressive microenvironment[10–12]. These findings support the hypothesis that the spontaneous adaptive immune response to premalignancy is a mechanism controlling PML-progression and that this might be therapeutically exploited for immunoprevention of cancer.

T cells are able to recognize aberrant peptides, including neoepitopes that are presented by tumor cells as a result of mutational events acquired during cancer initiation and progression. This endogenous T cell response to malignancy can be harnessed to mediate tumor regression using adoptive cell therapies[13] or through immune checkpoint inhibition[14]. Studies of non-small cell lung cancer (NSCLC) have shown that the T cell receptor (TCR) repertoire of NSCLC associates with the somatic genomic landscape[15]. The TCR repertoire of NSCLC has also been suggested as a biomarker for predicting the response to immune checkpoint blockade[16].

Previous studies have demonstrated that the TCR repertoire can be partially assembled from bulk RNA-sequencing (RNA-seq) data[17,18]. To enhance our understanding of the immune response to squamous PMLs and the interplay between the adaptive immune response and progression of PMLs, we characterized, using bulk RNA-seq data, the T-cell repertoire of squamous cell PMLs and its association with transcriptomic and clinical features. Our results indicate that regressive proliferative lesions have increased TCR repertoire clonality compared with persistent/progression PMLs.

## Methods

### Subject Population and Sample Collection and Processing

Endobronchial biopsies and brushings from high-risk subjects, undergoing lung cancer screening at approximately 1-year intervals via auto fluorescence bronchoscopy and CT, were profiled by RNA-Seq as previously described (n=295 biopsies, n=137 brushes, and n=50 patients) [9]. Briefly, endobronchial biopsies were obtained from abnormal fluorescing airway epithelium and brushings were obtained from normal appearing mainstem bronchi. RNA was extracted and sequenced, and the data was processed as previously described[9] to quantitate gene expression levels. The study was IRB approved and all participants consented to the study.

### TCR sequence assembly

We applied the TCR Repertoire Utilities for Solid Tissue/Tumor (TRUST, version 3.0) tool[17] to identify T-cell receptor (TCR) CDR3 sequences in the RNA-Seq data, applying it in single end mode to reads aligned to hg19 via STAR[19] (v. 3.0.0) (bam files from our previously published work[9]). TCR sequences with less than 6 amino acids were excluded from the analysis. We also compared the performance of TRUST to MiXCR[20] (version 2.1.10, using FASTQ files from our prior work[9]) in assembling CDR3 sequences. TCR sequences were assembled from both biopsy and brush samples, although only biopsies were used for diversity and TCR sharing and clonal expansion analyses. Eleven samples underwent targeted RNA TCR sequencing (IRepertoire, 2 replicates/sample) of the TCR alpha, beta, gamma and delta chains to compare TCR identification and TCR diversity metrics with those derived using RNA-seq data via TRUST (**Supplemental Table 1**).

### TCR repertoire diversity in PML biopsies

In the bulk RNA-Seq data, TCR diversity was measured as the number of unique clonotypes per 1,000 TCR reads, previously described as clonotypes per kilo-reads (CPK) [18]. CPK was chosen over Shannon entropy because it was not strongly correlated with total TCR reads in the bulk RNA-Seq data. For the targeted TCR sequencing data, where we achieved much higher TCR-read depth, diversity was measured as Shannon entropy. Spearman correlation was used to compare total TCR reads and TCR diversity between samples where TCR repertoires were measured by bulk RNA-seq and targeted TCR sequencing.

We used Spearman correlation to evaluate the association between CPK and the expression of an immune-associated geneset we previously identified and found to be associated with squamous PML regression[9]. The association between CPK and PML progression/persistence versus regression was evaluated with a two-sided Mann–Whitney U test. Correlation between CPK and the immune-associated gene-set was evaluated with Spearman correlation.

### TCR repertoire sharing and clonal expansion in PML biopsies

We compared the overlap of TCR repertoires in biopsies 1) between PMLs from different subjects, 2) within the same subject between PMLs acquired during the same procedure at different anatomic locations, and 3) within the same subject between PMLs acquired at the same anatomic location across different procedures. Fisher’s exact test was used to identify pairs of samples enriched in shared TCRs, comparing the number of CDR3 sequences shared and unique, against a background of all 39,303 TCRs. The mean odds ratios of these Fisher’s exact tests grouped by shared location, time, and subject were compared using the Tukey test.

To identify TCRs undergoing clonal expansion, we normalized counts of all TCRs to counts per thousand reads within each sample and defined expanded TCRs as those with normalized counts more than three standard deviations greater than the mean calculated across all samples. Samples with fewer than 250 reads total were removed from this analysis, as low total counts in a sample resulted in inflated normalized counts for individual TCRs. With a threshold of 250 reads, there was no longer a significant linear association between total reads and number of expanded clones (p>0.05 Pearson).

In order to identify whether the same TCR sequences were repeatedly expanded more often than expected by chance, we conducted a permutation test. We shuffled “expanded” vs “not expanded” labels of TCRs present 1000 times, then compared how often the same TCR sequence was expanded at multiple timepoints at each location within each patient.

### Identification of public and private TCRs

TCRs from both biopsies and brushes were defined as public if they were found in more than one patient or, for TCR beta, if they were also found in the peripheral blood of a previously described cohort of 666 healthy donors[21]. They were otherwise defined as private. While most of this work focused on biopsy samples, brushes were included in this analysis in order to more accurately identify public sequences. We also quantified the proportion of private and public that were found in publicly available databases, McPAS TCR[22] and VDJdb[23], which contain antigen-specific TCR sequences (both databases accessed 08/22/2019). In order to determine if more public versus private TCRs overlapped with TCR sequences found in the queried databases we used Fisher’s exact test.

## Results

### TCRs can be accurately assembled from bulk RNA-seq data in airway biopsies and brushes

We detected a total of 39,303 unique TCR sequences of at least 6 amino acids in length using the TRUST tool and RNA-seq from 294 biopsies of PMLs (1 biopsy did not generate TCR sequences) and 137 brushings of normal appearing airway epithelium from 50 subjects at high-risk for lung cancer. The TRUST TCR sequences were used for further analyses as MiXCR assembled fewer sequences (n=31,208 unique TCR sequences) and the sequences from both methods were highly overlapping (n=17,751 (45%) TCRs were shared, p < 1.5e-21 by hypergeometric test). Eleven representative biopsy samples underwent targeted RNA sequencing of the TCR loci, yielding an average of 585,589 TCR reads per sample (range 29,076-1,489,423) and 1,618 unique CDR3 sequences per sample (range 462-2,772). Both the total number of TCR reads identified and TCR diversity were significantly correlated across bulk and targeted sequencing results (r^2^ > .4, p < 0.001 both, **Supplemental Figure 1**). The TCR sequences from the bulk RNA-seq data and the targeted TCR-seq data were highly overlapping with an average of 21.9% of TCRs identified from bulk RNA-seq also being identified in targeted sequencing of the same sample, compared to 5.4% of bulk TCRs being identified in targeted sequencing from different samples (p = 6.05 x10-6, Mann-Whitney U test). Additionally, the percent of shared TCR sequences between the targeted sequencing and bulk samples was as least as high as the percent of shared TCR sequences between replicates of targeted sequencing, regardless of the method used to assemble the TCR sequences (**Supplemental Figure 2**). Based on these results, the TCR sequences assembled from the bulk RNA-seq data are representative of TCR sequences obtained from targeted TCR RNA sequencing.

### TCR diversity is associated with progression/persistence in proliferative PML biopsies

We quantified TCR diversity using CPK because it was least correlated with total TCR reads (Spearman’s rho=−0.26, p<0.0001) in comparison to other commonly used TCR diversity metrics (e.g. Shannon entropy, Simpson diversity index), (Spearman’s rho>0.8, p<2.2e-16) (**Supplemental Figure 3**). In our prior work[9], we identified modules of gene co-expression that we used to define PML molecular subtypes. Two of these nine modules are enriched for genes involved with immune functions. We found CPK to be negatively correlated with one of the immune modules that is associated with interferon signaling and antigen processing and presentation (Spearman’s rho = −0.34, p = 2.818e-09, **Figure 2A**). In other words, decreased TCR diversity is associated with higher expression of genes in this interferon-related immune module across all biopsies. Interestingly, we previously reported that high expression of the genes in this module is associated with regression within PMLs in the previously defined proliferative molecular subtype that is enriched for bronchial dysplasia. Here, we find that TCR diversity (CPK) was significantly decreased in the proliferative PMLs that regressed vs. PMLs that persisted/progressed (28.97 vs 35.57, p=0.045, Wilcoxon Rank Sum Test, n=50 samples; **Figure 2B**). These results indicate that increased clonality may be important in lesion regression.

**Figure 1.**
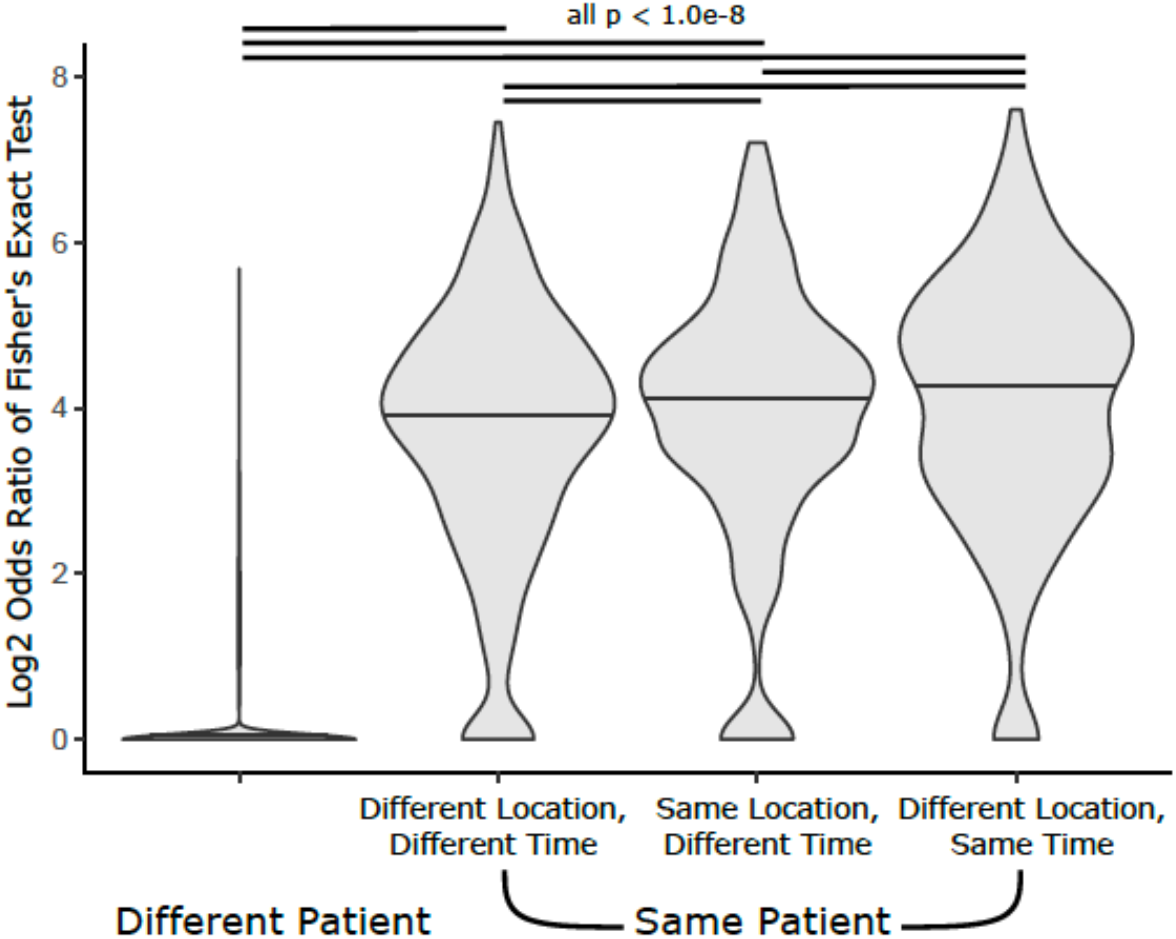
Enrichment of shared CDR3 sequences between pairs of samples. Log2 odds ratios of Fisher’s exact test of every pair of samples, grouped by shared location, time, and patient (ANOVA p < 2e-16). Significant differences between means is indicated by black lines (Tukey post-hoc FDR < 1.0e-8).

**Figure 2.**
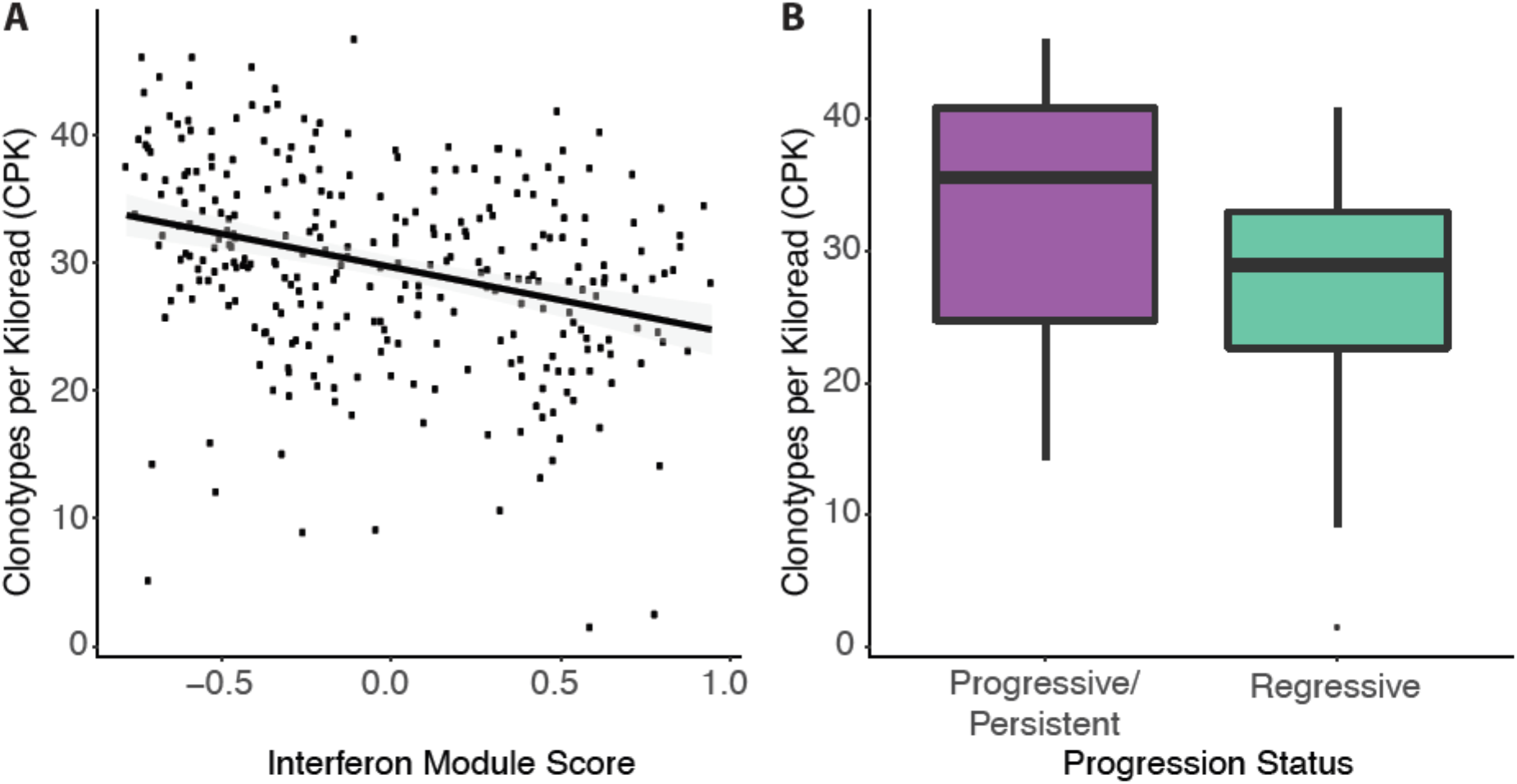
TCR clonality is associated with immune processes and premalignant lesion outcome. **(A**.) Clonotypes per Kiloread (CPK) is negatively correlated with a gene expression module of interferon response previously shown to be negatively associated with lesion progression (Spearman’s rho −0.34, p = 2.818e-09). (**B**.) Clonotypes per kiloread (CPK) in *Proliferative* lesions, by progression status (progressive/persistent lesions, purple; regressive lesions, turquoise). Progressive lesions have significantly higher CPK than regressing lesions (p = 0.045, n=22 progressive/persistent and n=28 regressive samples).

### TCR clone sharing and clonal expansion is increased within patient PMLs

On average across the subjects, endobronchial biopsies were obtained from different anatomic locations during multiple screening bronchoscopies. Out of the 34,251 CDR3 sequences identified in biopsy samples, 2,494 (7.3%) were present in more than one biopsy in our cohort. In order to identify patterns in CDR3 sharing between biopsies, we identified enrichment of shared CDR3 sequences for all possible pairs of PML biopsies using Fisher’s exact test. Only 0.01% of pairs of samples from different patients showed significant enrichment (at FDR < 0.01) of shared CDR3 sequences. Among samples from the same patients, we detected higher TCR clone sharing among samples found at different anatomic locations at identical time points (61.9% of pairs enriched at FDR <0.01, mean OR = 26.8) and among samples taken from the same anatomic location at separate time points (55.0% enriched at FDR <0.01, mean OR = 23.5) versus samples at different locations and time points (50% enriched at FDR<0.01, mean OR = 20.3) (**Table 1)**. ANOVA with Tukey’s post-hoc (adj. p<0.01, **Figure 1**).

**Table 1.** Shared sequences between samples. Percent of pairs of samples enriched for shared sequences, grouped by shared location, shared time, and shared patient. Enrichment was determined by a Fisher’s exact test, FDR cutoff of 0.01 and Odds ratio > 1.

| Factors Shared |  |  | Non-enriched Pairs | Enriched Pairs | Percent Enriched | Mean Odds Ratio |
| --- | --- | --- | --- | --- | --- | --- |
| Patient | Location | Time |  |  |  |  |
|  |  |  | 83,004 | 8 | 0.009% | 0.214 |
| X |  |  | 736 | 746 | 50.3% | 3.69 |
| X | X |  | 206 | 252 | 55.0% | 3.90 |
| X |  | X | 230 | 374 | 61.9% | 4.09 |

Among the 74 lesions with at least 250 total TCR counts, we identified 344 TCRs likely to have undergone clonal expansion in PML biopsies based on increased normalized abundance three standard deviations above the mean across all samples. Among TCRs that were only found in a single sample, 3.1% of TCRs were expanded, significantly less than in TCRs that were identified in multiple samples within the same patient (10.3%, p < 0.001; OR = 3.607 Fisher’s Test). Furthermore, we identified 10 TCRs that were expanded at multiple time points in the same lesion at the same location, more than would be expected if all TCRs recurring in a patient were equally likely to be expanded (p<0.001). The result indicates that TCRs can persist in an expanded state for multiple years in the same lesion.

### Public TCRs commonly target infectious antigens

Among all TCRs identified in both brushes and biopsies, 4,543 (11.6%) were considered public, meaning shared between multiple patients in our cohort or, for TCR beta sequences, identified in a previously described cohort of 666 people[21]. The proportion of public TCRs was higher among clones expanded in biopsies, at 26.1% (Fisher’s p = 1.1e-10, OR 2.36). Public TCRs were more commonly found in the VDJdb and McPAS TCR databases than non-public (private) TCRs (8.69% and 1.53%, p < 0.001, Fisher’s exact test, OR 6.12). The VDJdb and McPAS databases predominantly annotate TCRs targeting infectious antigens. TCRs targeting common viral pathogens, including predominantly influenza, cytomegalovirus and Epstein Barr Virus, but also Yellow Fever, Hepatitis C and Human Immunodeficiency Virus, were detected. Few TCRs were annotated as recognizing Bone Marrow Stromal Antigen 2 (BST2) and Melanoma Antigen Recognized by T cells 1 (MART-1). As expected, the identification of a public TCR is more likely to represent a TCR targeting an infectious antigen than a cancer-specific antigen, although our analysis is limited by the sequences annotated in the VDJdb and McPAS databases.

## Discussion

Lung squamous cell carcinoma arises in the epithelial layer of the bronchial airways and is often preceded by the development of bronchial premalignant lesions (PMLs). Bronchial PMLs have variable outcome and biomarkers of PML progression and therapies to intercept lung cancer development at this early stage are needed. We have previously reported the identification of four molecular subtypes of PMLs based on bulk RNA-seq profiling of endobronchial biopsies and brushes from high-risk smokers undergoing lung cancer screening[9]. The proliferative subtype was enriched with bronchial dysplasia and progressive/persistent PMLs in this subtype showed decreased expression of interferon signaling and antigen processing/presentation pathways compared with regressive lesions. In this study, we leverage the bulk RNA-seq data from this prior work to characterize the TCR repertoire of lung squamous PMLs and its association with PML progression.

Given that bulk RNA sequencing is a relatively insensitive method of characterizing the TCR repertoire, capturing only a fraction of the TCRs, we confirmed on a subset of samples that total TCR reads and TCR diversity was significantly correlated between TCRs derived from bulk RNA-seq versus targeted TCR-seq data. Using the TCRs derived from bulk RNA-seq and the gene expression data, we found that in the biopsies, a decrease in TCR diversity or an increase in TCR clonal selection is significantly associated with a metagene score for the gene signature associated with interferon and antigen processing and presentation pathways. Interestingly, we also found increased TCR clonal selection in regressive versus progressive/persistent lesions belonging to the proliferative subtype. These results suggest that regressive high-grade lesions are recognizing neoantigens, and further single T cell sequencing and functional studies are needed to evaluate the utility of these for immunoprevention of lung squamous cell carcinoma.

Since the biopsies in this study were serially collected at different anatomic locations in the lung, we were also able to quantify the amount of TCR clone sharing between or within patients as a function of space and time. We detected a high degree of clone sharing between biopsies from the same patients, and the degree of sharing was higher for samples obtained from the same time point versus the same anatomic location at separate time points. Shared TCRs within a patient were more likely to be expanded than TCRs found in a single sample. We observed 10 TCRs that were expanded at multiple time points in the same lesion at the same location suggesting the potential existence of lesion-specific neoantigens. Additionally, our results show that TCR repertoires significantly overlap in different areas of the airway and suggest that a field immunity effect may also exist concurrently with lesion-specific immunity. This finding parallels our prior work showing field-level transcriptomic changes associated with PML and lung cancer development in distant, normal-appearing tissue from the nasal turbinate and the mainstem bronchus[24–26]. Currently, however, we do not have evidence that these shared or expanded TCR clones are targeting PML-related antigens and our results are challenged by limited sequencing depth and nonuniform sampling of the lung. In the future, TCRs detected in regressing lesions that may mediate PML regression need to be functionally characterized. Additionally, further comprehensive characterization of ubiquitous and regional TCR repertoires in the context of lung preneoplastic lesions is needed as has been done for NSCLC [15,27].

In order to further characterize the TCR repertoire, we classified TCRs as public (found in more than one patient) and private (found in only one patient). Public TCRs were more likely to be clonally expanded than TCR sequences found only in one patient. Additionally, they were more likely to have known infectious antigen specificity in our dataset, consistent with previous evidence of shared TCRs in response to infectious agents[21]. The enrichment of TCRs with known antigenic specificities among the public TCR clones could be explained by the preponderance of infectious epitopes in the existing databases of TCR sequences and their antigenic specificity. TCRs targeting tumor neoantigens are less likely to be shared among patients because individual patients and different PMLs carry distinct mutations that can be presented in differing Human Leukocyte Antigen (HLA) contexts. There are exceptions, however, as public TCRs that target a single neoantigen have been demonstrated in response to the common *KRAS* exon 12 G12D mutation in colorectal cancer, and can be leveraged for adoptive T-cell therapy[28]. In future studies, it will be important to distinguish between immune responses to infections and cancer development and to identify neoantigen recognition in specific HLA contexts as this may be relevant for immunoprevention[29].

## Conclusion

Our study provides TCR characterization of bronchial lesions that are precursors of lung squamous cell carcinoma suggesting that TCR diversity measured via RNA may help predict the host immune response at the earliest stages of cancer development and may, in the future, aid in the development of immunoprevention strategies for lung cancer.

## Data Availability

RNA sequencing data from human endobronchial biopsies and brushings has been previously deposited in the NCBI Gene Expression Omnibus under accession code GSE109743.

http://www.ncbi.nlm.nih.gov/geo/query/acc.cgi?acc=GSE109743

## List of abbreviations

PML: premalignant lesion
CPK: clonotypes per kilobase
TCR: T cell receptor
PCGA: Pre-Cancer Genome Atlas

## Declarations

### Ethics approval and consent to participate

The Institutional Review Boards at Boston University Medical Center and Roswell Comprehensive Cancer Center approved the study and all subjects provided written informed consent.

### Availability of data and material

RNA sequencing data from human endobronchial biopsies and brushings has been previously deposited in the NCBI Gene Expression Omnibus under accession code GSE109743 [http://www.ncbi.nlm.nih.gov/geo/query/acc.cgi?acc=GSE109743].

### Authors’ contributions

Study conception and design (JEB, AM, MEL); collection of clinical samples (MER); sample processing for bulk RNA sequencing (GL, SZ, HL); sample and clinical data management (AP, ACG); bulk RNA sequencing pipeline to generate alignments and gene expression (YK, JDC); TCR read generation and data analyses (AM, CM, JEB); writing of the manuscript (JEB, AM, CM); editing of the manuscript (JEB, AM, CM, JDC, SAM, MEL, AES).

## Acknowledgements

We would like to thank David L. Coleman MD, Gopal K. Yadavalli MD and Ashish Upadhyay MD from Boston University for their mentorship and support for AM.

