## Supplemental Figure 1 for "Elevated T cell repertoire diversity is associated with progression of lung squamous cell premalignant lesions"

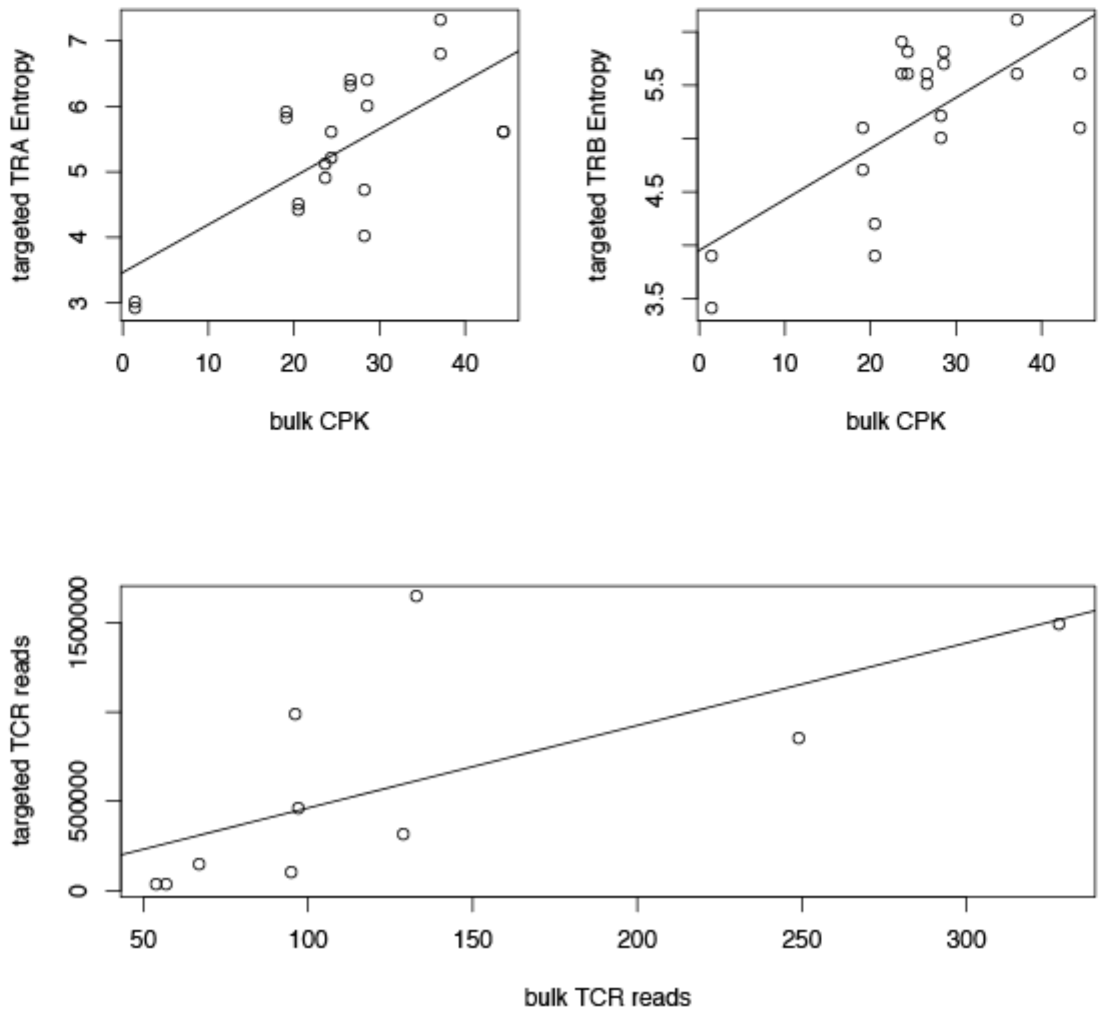


**Supplementary Figure 1. TCR diversity is correlated between metrics derived from bulk RNA sequencing data and targeted TCR data.** The clonotypes per kiloread (CPK) metric derived from the bulk RNA sequencing data correlates well with targeted TCR-seq metrics of entropy, in both TRA **(A.)** and TRB **(B.)** receptors ($p < 1.0e-3$, both). Correlations could not be computed for TRD and TRG reads because few reads were detected in the bulk RNA sequencing. (**C.)** Number of TCR reads total also correlated significantly between targeted and bulk RNA sequencing ($p = 0.029$).
