## Supplemental Figure 2 for "Elevated T cell repertoire diversity is associated with progression of lung squamous cell premalignant lesions"

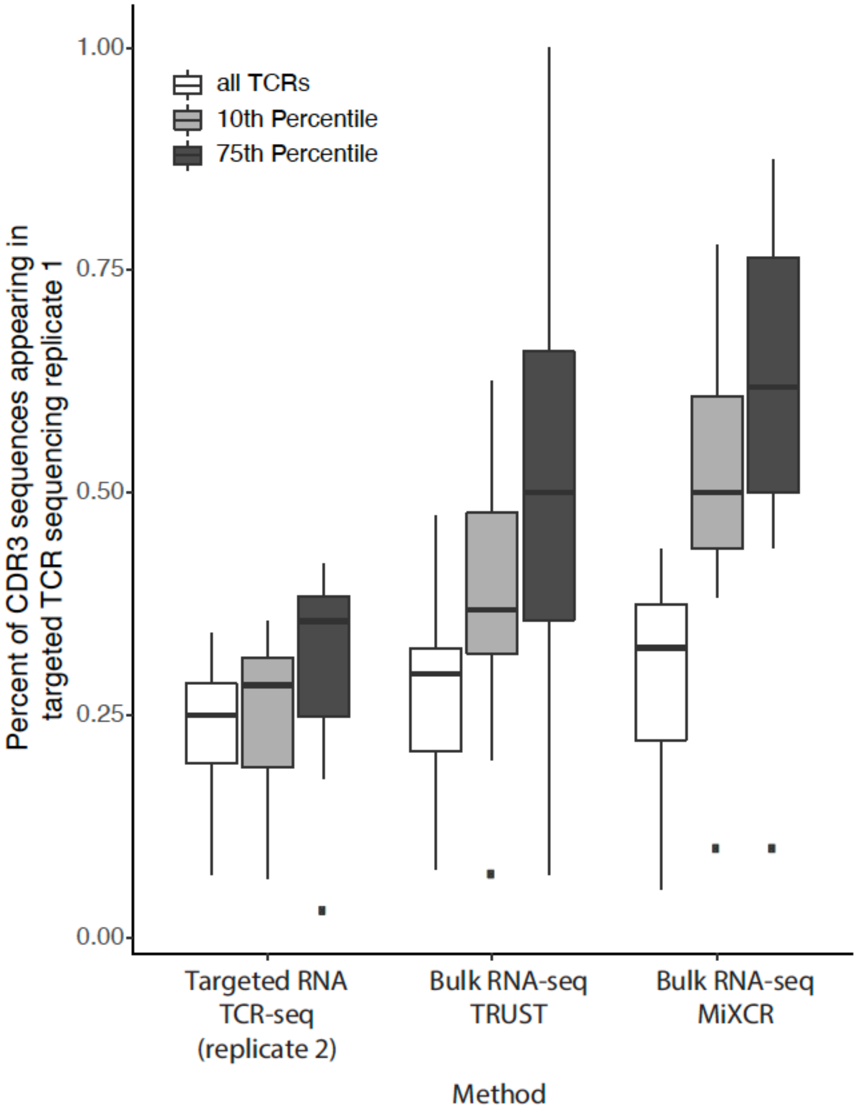


**Supplementary Figure 2. Shared CDR3 amino acid sequences from bulk RNA-seq assembled TCRs and targeted RNA TCR-seq TCRs.** The targeted RNA TCR sequencing generated data for 2 replicates per sample (n=11 samples). The boxplot shows the percent of CDR3 amino acid sequences from each method (Targeted RNA TCR-seq Replicate 2, Bulk RNA-seq TRUST, or Bulk RNA-seq MiXCR) that was also found in the targeted TCR-seq Replicate 1. Across all TCRs, no difference is found in the percent of CDR3 sequences from each group that appear in the replicate (Kruskal Wallace p>0.1). Percentages of shared CDR3 sequences were plotted for all TCR sequences (white boxes) or TCR sequences in at least the 10$^{th}$ (gray boxes) or 75$^{th}$ (dark gray boxes) percentiles of abundance for each sample in each method. When applying increasing thresholds of TCR abundance, all methods showed increased agreement with the TCR-seq replicate.
