## Supplemental Figure 3 for "Elevated T cell repertoire diversity is associated with progression of lung squamous cell premalignant lesions"

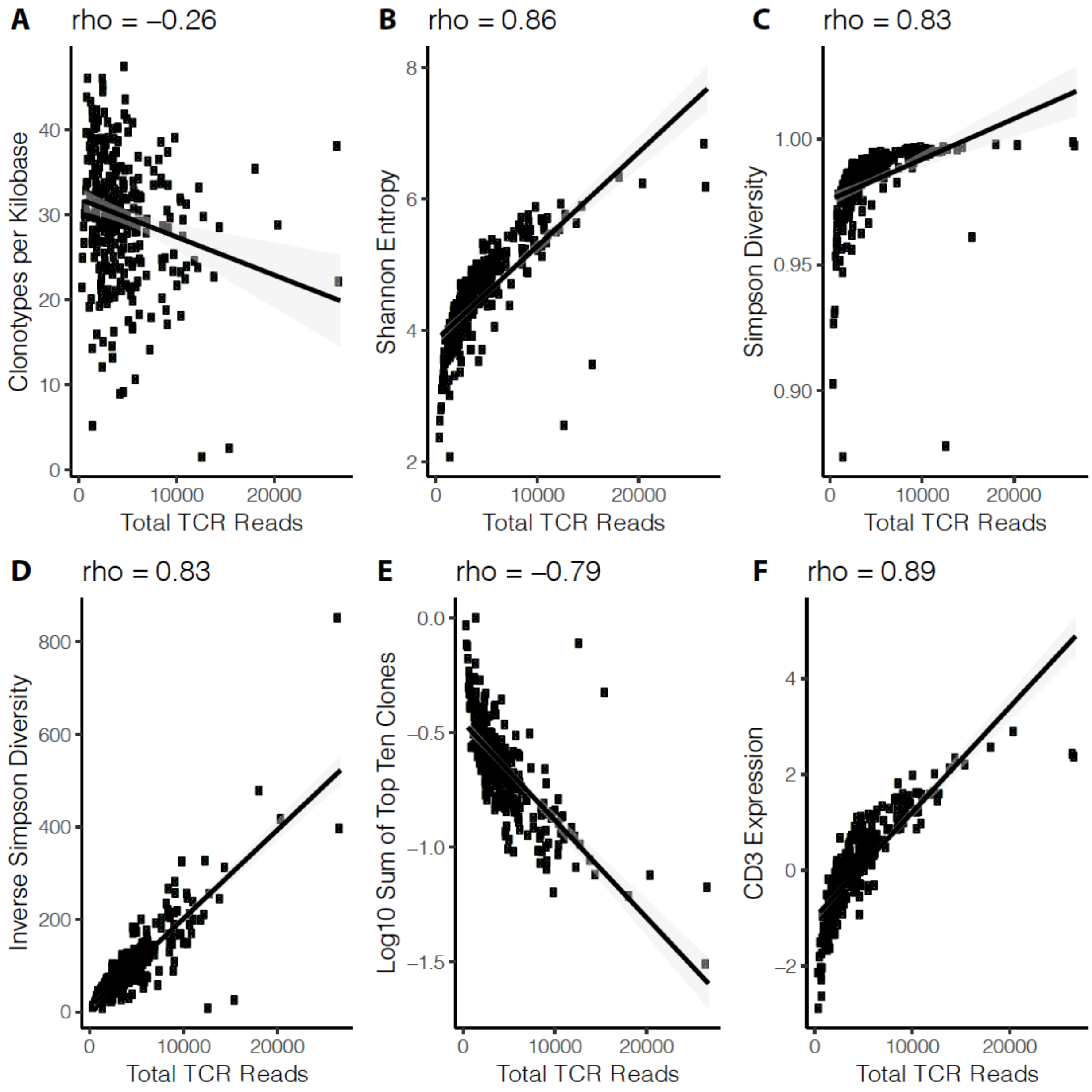


**Supplementary Figure 3. Diversity metrics correlated with total number of TCR reads**. The plots show the correlation between total TCR reads and diversity metrics and T cell gene expression. The plots are as follows: **(A.)** clonotypes per kiloread (CPK), **(B.)** Shannon entropy, **(C.)** Simpson diversity, **(D.)** inverse Simpson diversity, **(E.)** log10 sum of the top 10 clones, and **(F.)** CD3 gene expression. Clonotypes per kiloread (CPK) was the diversity metric that demonstrated the weakest correlation to total TCR reads.
