## Supplemental Table 1 for "Elevated T cell repertoire diversity is associated with progression of lung squamous cell premalignant lesions"

| PCGA ID | Chain | Total Reads | Unique CDR3 Reads |
|---|---|---|---|
| PCGA-01-0001-037-19118-00482BX-1007555R | TRA | 35743 | 230 |
| PCGA-01-0001-037-19118-00482BX-1007555R | TRA | 33153 | 234 |
| PCGA-01-0001-037-19118-00482BX-1007555R | TRB | 12548 | 119 |
| PCGA-01-0001-037-19118-00482BX-1007555R | TRB | 11306 | 123 |
| PCGA-01-0001-037-19118-00482BX-1007555R | TRD | 734 | 10 |
| PCGA-01-0001-037-19118-00482BX-1007555R | TRD | 394 | 4 |
| PCGA-01-0001-037-19118-00482BX-1007555R | TRG | 3642 | 28 |
| PCGA-01-0001-037-19118-00482BX-1007555R | TRG | 2149 | 24 |
| PCGA-01-0001-037-19762-00898BX-1007486R | TRA | 470869 | 740 |
| PCGA-01-0001-037-19762-00898BX-1007486R | TRA | 461677 | 746 |
| PCGA-01-0001-037-19762-00898BX-1007486R | TRB | 147864 | 389 |
| PCGA-01-0001-037-19762-00898BX-1007486R | TRB | 170537 | 444 |
| PCGA-01-0001-037-19762-00898BX-1007486R | TRD | 31061 | 92 |
| PCGA-01-0001-037-19762-00898BX-1007486R | TRD | 40305 | 97 |
| PCGA-01-0001-037-19762-00898BX-1007486R | TRG | 81779 | 112 |
| PCGA-01-0001-037-19762-00898BX-1007486R | TRG | 85331 | 152 |
| PCGA-01-0001-037-20126-01120BX-1031613R | TRA | 124142 | 446 |
| PCGA-01-0001-037-20126-01120BX-1031613R | TRA | 110040 | 421 |
| PCGA-01-0001-037-20126-01120BX-1031613R | TRB | 69057 | 362 |
| PCGA-01-0001-037-20126-01120BX-1031613R | TRB | 58152 | 348 |
| PCGA-01-0001-037-20126-01120BX-1031613R | TRD | 7978 | 44 |
| PCGA-01-0001-037-20126-01120BX-1031613R | TRD | 7768 | 54 |
| PCGA-01-0001-037-20126-01120BX-1031613R | TRG | 11734 | 65 |
| PCGA-01-0001-037-20126-01120BX-1031613R | TRG | 11553 | 65 |
| PCGA-01-0001-070-19300-00587BX-1007583R | TRA | 125474 | 244 |
| PCGA-01-0001-070-19300-00587BX-1007583R | TRA | 136958 | 239 |
| PCGA-01-0001-070-19300-00587BX-1007583R | TRB | 66573 | 209 |
| PCGA-01-0001-070-19300-00587BX-1007583R | TRB | 40470 | 117 |
| PCGA-01-0001-070-19300-00587BX-1007583R | TRD | 21009 | 56 |
| PCGA-01-0001-070-19300-00587BX-1007583R | TRD | 12532 | 50 |
| PCGA-01-0001-070-19300-00587BX-1007583R | TRG | 22739 | 66 |
| PCGA-01-0001-070-19300-00587BX-1007583R | TRG | 28001 | 39 |
| PCGA-01-0001-089-20126-01118BX-1031611R | TRA | 263802 | 486 |
| PCGA-01-0001-089-20126-01118BX-1031611R | TRA | 245170 | 538 |
| PCGA-01-0001-089-20126-01118BX-1031611R | TRB | 119886 | 433 |
| PCGA-01-0001-089-20126-01118BX-1031611R | TRB | 90949 | 361 |
| PCGA-01-0001-089-20126-01118BX-1031611R | TRD | 30969 | 76 |
| PCGA-01-0001-089-20126-01118BX-1031611R | TRD | 39131 | 109 |
| PCGA-01-0001-089-20126-01118BX-1031611R | TRG | 21206 | 64 |
| PCGA-01-0001-089-20126-01118BX-1031611R | TRG | 39372 | 95 |
| PCGA-01-0021-025-20449-01091BX-1031600R | TRA | 7903 | 114 |
| PCGA-01-0021-025-20449-01091BX-1031600R | TRA | 8879 | 131 |
| PCGA-01-0021-025-20449-01091BX-1031600R | TRB | 5379 | 89 |
| PCGA-01-0021-025-20449-01091BX-1031600R | TRB | 5127 | 85 |
| PCGA-01-0021-025-20449-01091BX-1031600R | TRD | 178 | 3 |
| PCGA-01-0021-025-20449-01091BX-1031600R | TRD | 73 | 2 |
| PCGA-01-0021-025-20449-01091BX-1031600R | TRG | 648 | 17 |
| PCGA-01-0021-025-20449-01091BX-1031600R | TRG | 889 | 21 |
| PCGA-01-0021-050-19434-00465BX-1007550R | TRA | 103665 | 467 |
| PCGA-01-0021-050-19434-00465BX-1007550R | TRA | 97156 | 523 |
| PCGA-01-0021-050-19434-00465BX-1007550R | TRB | 48015 | 265 |
| PCGA-01-0021-050-19434-00465BX-1007550R | TRB | 40966 | 268 |
| PCGA-01-0021-050-19434-00465BX-1007550R | TRD | 4168 | 37 |
| PCGA-01-0021-050-19434-00465BX-1007550R | TRD | 1700 | 17 |
| PCGA-01-0021-050-19434-00465BX-1007550R | TRG | 15009 | 77 |
| PCGA-01-0021-050-19434-00465BX-1007550R | TRG | 8311 | 62 |
| PCGA-01-0021-070-20449-01089BX-1031598R | TRA | 361340 | 505 |
| PCGA-01-0021-070-20449-01089BX-1031598R | TRA | 569838 | 595 |
| PCGA-01-0021-070-20449-01089BX-1031598R | TRB | 304895 | 630 |
| PCGA-01-0021-070-20449-01089BX-1031598R | TRB | 332628 | 474 |
| PCGA-01-0021-070-20449-01089BX-1031598R | TRD | 10354 | 28 |
| PCGA-01-0021-070-20449-01089BX-1031598R | TRD | 28781 | 37 |
| PCGA-01-0021-070-20449-01089BX-1031598R | TRG | 25702 | 48 |
| PCGA-01-0021-070-20449-01089BX-1031598R | TRG | 8122 | 25 |
| PCGA-01-0042-010-23911-01028BX-1033960R | TRA | 217964 | 309 |
| PCGA-01-0042-010-23911-01028BX-1033960R | TRA | 304970 | 543 |
| PCGA-01-0042-010-23911-01028BX-1033960R | TRB | 219443 | 599 |
| PCGA-01-0042-010-23911-01028BX-1033960R | TRB | 174843 | 625 |
| PCGA-01-0042-010-23911-01028BX-1033960R | TRD | 1297 | 12 |
| PCGA-01-0042-010-23911-01028BX-1033960R | TRG | 32978 | 101 |
| PCGA-01-0042-010-23911-01028BX-1033960R | TRG | 31082 | 85 |
| PCGA-01-0042-048-23680-00907BX-1033955R | TRA | 9401 | 277 |
| PCGA-01-0042-048-23680-00907BX-1033955R | TRA | 9178 | 290 |
| PCGA-01-0042-048-23680-00907BX-1033955R | TRB | 7906 | 322 |
| PCGA-01-0042-048-23680-00907BX-1033955R | TRB | 6621 | 330 |
| PCGA-01-0042-048-23680-00907BX-1033955R | TRD | 15 | 3 |
| PCGA-01-0042-048-23680-00907BX-1033955R | TRD | 34 | 8 |
| PCGA-01-0042-048-23680-00907BX-1033955R | TRG | 877 | 32 |
| PCGA-01-0042-048-23680-00907BX-1033955R | TRG | 760 | 23 |
| PCGA-01-0042-048-23911-01029BX-1034033R | TRA | 39007 | 237 |
| PCGA-01-0042-048-23911-01029BX-1034033R | TRA | 42164 | 293 |
| PCGA-01-0042-048-23911-01029BX-1034033R | TRB | 27234 | 259 |
| PCGA-01-0042-048-23911-01029BX-1034033R | TRB | 24916 | 302 |
| PCGA-01-0042-048-23911-01029BX-1034033R | TRD | 92 | 3 |
| PCGA-01-0042-048-23911-01029BX-1034033R | TRD | 106 | 1 |
| PCGA-01-0042-048-23911-01029BX-1034033R | TRG | 3262 | 38 |
| PCGA-01-0042-048-23911-01029BX-1034033R | TRG | 3848 | 53 |

**Supplementary Table 1. Samples that underwent targeted RNA TCR sequencing.** For each sample, the chain sequenced, the total number of reads sequenced, and the number of unique CDR3 reads are listed.
